# Applications of ICG in Breast Cancer for Sentinel Lymph Node Mapping: A Scoping Review Protocol

**DOI:** 10.1101/2024.07.30.24311256

**Authors:** Feryal N. Kurdi, Yahya N. Kurdi, Igor V. Reshetov

## Abstract

**Objective:** The objective of this scoping review is to evaluate the current literature on the use of Indocyanine Green (ICG) in sentinel lymph node (SLN) mapping for breast cancer patients. This review aims to assess the accuracy, efficacy, and safety of ICG in this context and to identify gaps in the existing research. The outcomes will contribute to the development of further research as part of a PhD project.

**Introduction:** Breast cancer is the leading cause of morbidity and mortality worldwide. Accurate SLN mapping is crucial for staging and treatment planning in early-stage breast cancer. ICG has emerged as a promising agent for fluorescence imaging in SLN mapping. However, comprehensive assessment of its clinical utility, including accuracy and adverse effects, remains limited. This scoping review aims to consolidate evidence on the use of ICG in breast cancer SLN mapping.

**Inclusion criteria:** Patients with early-stage breast cancer (T1, T2), selected T3 cases where sentinel lymph node biopsy is accurate, and clinically node-negative breast cancer. The intervention includes studies using ICG for SLN mapping and assessment of fluorescence imaging cameras.

**Methods:** Five electronic databases will be searched (PubMed, EMBASE, MEDLINE, Web of Science, and SCOPUS) using search strategies developed in consultation with the academic supervisor. The search strategy is set to human studies published in English within the last 11 years. All retrieved citations will be imported to Zotero and then uploaded to Covidence for screening of titles, abstracts, and full text according to pre-specified inclusion criteria. Citations meeting the inclusion criteria for full-text review will have their data extracted by two independent reviewers, with disagreements resolved by discussion. A data extraction tool will be developed to capture full details about the participants, concept, and context, and findings relevant to the scoping review will be summarized.

## Introduction

Sentinel lymph node (SLN) biopsy plays a crucial role in staging and prognosis in breast cancer management. The SLN is the initial lymph node to which breast cancer cells are likely to metastasize, and the presence of cancer cells in the SLN indicates a higher likelihood of further metastasis to other lymph nodes and distant organs.^1^

SLN biopsy involves injecting a tracer substance into the breast, which migrates to the SLN. The SLN is then identified, excised, and examined for cancer cells. If the SLN is free of cancer cells, it suggests that the cancer has not spread to other lymph nodes, eliminating the need for additional lymph node dissection. Conversely, if the SLN contains metastases, further dissection is typically required.^2^

Over the past two decades, the use of SLN biopsy utilizing blue dye and radiotracers (BD-RI) has been established as the diagnostic standard of care for patients with early-stage breast cancer who have clinically negative lymph nodes.^3,4^

However, these methods come with certain drawbacks, including the potential for allergic reactions to the blue dye and the necessity of nuclear medicine facilities for radiotracer injection and detection. In a cohort undergoing blue dye and radiotracer injection (BD-RI) procedures, a small number of adverse reactions, such as skin tattooing and anaphylaxis, were reported.^5^

### Emergence of ICG Technology

In recent years, near-infrared (NIR) fluorescence imaging using indocyanine green (ICG) has emerged as an alternative approach for SLN mapping in breast cancer patients. ICG, a fluorescent dye, is injected into the breast and migrates to the SLNs. A NIR camera detects the fluorescence emitted by ICG, enabling the surgeon to identify and excise the SLNs. ^6^

This technology offers several advantages over traditional methods, including enhanced visualization of SLNs, a lower risk of allergic reactions, and the elimination of the need for nuclear medicine facilities. Furthermore, ICG has an excellent safety profile, with adverse events occurring at a low incidence rate of 1 in 42,000 patients.^7–10^

The importance of this topic stems from the potential of ICG technology to enhance the accuracy and safety of SLN mapping in breast cancer patients. Precise identification and removal of the SLN are crucial for accurate staging and prognosis. Inaccurate SLN identification can lead to unnecessary lymph node dissection, resulting in complications such as lymphedema and impaired arm function. Sampling a larger number of SLNs may increase the risk of upper limb lymphedema, sensory deficits, and reduced shoulder function.

Landmark trials have shown a significant difference in morbidity rates when comparing SLN biopsy to axillary dissection, with rates of 25% and 70%, respectively.^3,11^ Recent studies have reported excising, on average, two nodes per patient, likely due to advancements in NIR technology and ICG fluorescence protocols.^12–16^ Nevertheless, further research is essential to assess the long-term outcomes and cost-effectiveness of ICG technology compared to traditional methods.

In summary, the significance of SLN mapping using ICG technology in breast cancer lies in its potential to enhance accuracy and safety, reduce complications, and improve patient outcomes.^17^ Further studies are necessary to assess the long-term efficacy and cost-effectiveness of this technique and to identify the patient populations most likely to benefit.

### Assessing the extent of the current literature

Although ICG technology has been used for SLN mapping in breast cancer patients, initial searches in 2022 revealed limited data on the feasibility, safety, and effectiveness of this technique. At that time, a preliminary search of SCOPUS, MEDLINE, EMBASE, and PubMed identified no current or forthcoming systematic reviews or scoping reviews on the topic. However, recent searches indicate a substantial increase in research and reviews, reflecting growing interest and evidence in this area.

**The objective of this scoping review is to** assess the extent of the literature around the evaluation and integration of emergent evidence for benefits and about harms exploring the feasibility, safety, and effectiveness of SLN mapping using ICG technology in a larger cohort of breast cancer patients, and guidance for clinical decision making.

This scoping review could also identify specific patient populations, such as those with higher BMIs, who may benefit most from ICG technology. Additionally, patients who have undergone neoadjuvant therapy could be particularly advantageous candidates.

Factors such as the type of NIR cameras used, the learning curve for surgeons to become proficient with ICG for SLN detection, the availability of ICG and radioisotopes, the presence of nuclear medicine facilities, regional variations in ICG usage, and cost comparisons with the gold standard are also critical considerations in the broader adoption of this technology.

The outcomes of the scoping review will inform, and frame three subsequent pieces of work planned as part of a PhD as follows:

1. Prospective Cohort Study on the Long-term Outcomes of ICG in SLN mapping
2. Systematic Review and Meta-Analysis of ICG for SLN mapping in Breast Cancer
3. Development of Standardized Clinical Guidelines and Protocols for the Use of ICG in SLN mapping in Breast Cancer Patients

The Participants Concept Context (PCC) framework for this scoping review defines the:

▪ participants as patients with early-stage breast cancer
▪ concept as the use of ICG for SLN mapping in breast cancer patients
▪ context where SLN mapping is performed as part of breast cancer staging and treatment planning.

## Review questions

What do we know about the evaluation and integration of emergent evidence on the use of ICG for SLN mapping in breast cancer patients into clinical practice and decision-making?

I. **To what extent** is emergent evidence on the feasibility, safety, and effectiveness of ICG for SLN mapping integrated into clinical guidelines and decision-making processes?
II. **How** is emergent evidence on the use of ICG for SLN mapping evaluated and incorporated into clinical guidelines and decision-making processes?

For the purposes of this scoping review, emergent evidence refers to new research findings on ICG for SLN mapping that have emerged post-market and have not yet been fully integrated into clinical guidelines and practice.

## Eligibility criteria

### Participants

▪ Patients with early-stage breast cancer (T1, T2).
▪ Selected T3 cases where sentinel lymph node biopsy has been shown to be accurate.

### Concept

▪ The use of ICG for sentinel lymph node mapping in breast cancer patients.
▪ Assessment of imaging techniques and devices used in conjunction with ICG for SLN mapping.

### Context

▪ Clinical settings where SLN mapping is performed as part of breast cancer staging and treatment planning.

#### Types of Sources

This scoping review will consider both experimental and quasi-experimental study designs including controlled before and after studies and controlled interrupted time-series studies. In addition, analytical observational studies including prospective and retrospective cohort studies, case-control studies and analytical cross-sectional studies will be considered for inclusion. This review will also consider descriptive observational study designs such as descriptive cross-sectional studies for inclusion.

Qualitative studies that focus on qualitative data will be considered for inclusion.

## Methods

The proposed scoping review will be guided by the JBI methodology for scoping reviews. ^18^

### Search strategy

The search strategy aims to locate both published and unpublished articles. An initial limited search of PubMed, EMBASE, MEDLINE, Web of Science, and SCOPUS was undertaken to identify relevant articles on the use of ICG for SLN mapping in breast cancer. In consultation with an academic supervisor, the text words in the titles and abstracts of relevant articles, and the index terms used to describe these articles were used to develop a comprehensive search strategy for PubMed, EMBASE, MEDLINE, Web of Science, and SCOPUS (see Appendix 1). This strategy, including all identified keywords and index terms, will be adapted for each included database. The articles sourced from all included sources of evidence will be exported into Zotero (www.zotero.org).

Only articles published in English will be included due to the language proficiency of the reviewers. Articles published since January 1, 2014, will be included to ensure relevance, aligning with the project’s consideration of recent data and the ongoing advancements in SLN mapping techniques using ICG.

### Study/Source of Evidence selection

Following the search, all identified articles will be exported into Zotero. Then the remaining articles will be uploaded into Covidence (www.covidence.org). Titles and abstracts will then be screened by the lead author (KF) against the inclusion criteria for the scoping review. Potentially relevant articles will be retrieved in full and included in Covidence. The full-text of these articles will be assessed in detail against the inclusion criteria by two independent reviewers, including (KF). Reasons for exclusion of sources of evidence at the full-text stage that do not meet the inclusion criteria will be recorded and reported in the scoping review. Any disagreements that arise between the reviewers at each stage of the selection process will be resolved through discussion, or with an additional reviewer. The results of the search and the study inclusion process will be reported in full in the final scoping review and presented in a Preferred Reporting Items for Systematic Reviews and Meta-analyses extension for scoping review as extracted from Covidence.

### Data Extraction

Data will be extracted from all articles included in the scoping review by two independent reviewers (including KF) using a data extraction tool developed by the lead reviewer (KF) and piloted with about 15 articles to refine and improve it. The data extracted will include specific details about the participants, concept, context, study methods and key findings relevant to the scoping review questions and will be imported into either Covidence or Excel.

A draft extraction form is provided (see Appendix 2). The draft data extraction tool will be modified and revised as necessary during the process of extracting data from each included article. Modifications will be detailed in the scoping review. Any disagreements that arise between the reviewers will be resolved through discussion, or with an additional reviewer. If appropriate, authors of articles will be contacted to request missing or additional data, where required.

### Data Analysis and Presentation

The evidence presented will directly respond to the scoping review objective and questions. The data will be presented graphically or in diagrammatic or tabular form. A narrative summary will accompany the tabulated and/or charted results and will describe how the results relate to the reviews objective and questions.

## Data Availability

All data produced in the present study are available upon reasonable request to the authors

## Acknowledgements

This scoping review is to contribute in part to a Doctor of Philosophy degree for (KF).

## Data and Software Availability

No data are associated with this article

## Appendices

### Appendix 1: Search strategy

Search strategies will be developed in consultation with an academic supervisor for PubMed, EMBASE, MEDLINE, Web of Science, and SCOPUS. All search strategies will be available on request.

Database: Embase

(‘indocyanine green’/exp OR ‘indocyanine green’ OR

((‘indocyanine’/exp OR indocyanine) AND (‘green’/exp OR green))

OR ‘icg’/exp OR icg OR ‘fluorescence imaging’/exp OR ‘fluorescence imaging’

OR ((‘fluorescence’/exp OR fluorescence) AND (‘imaging’/exp OR imaging)))

AND

(‘sentinel lymph node mapping’/exp OR ‘sentinel lymph node mapping’ OR

((‘sentinel’/exp OR sentinel) AND (‘lymph’/exp OR lymph) AND node

AND (‘mapping’/exp OR mapping)) OR ‘sentinel lymph node biopsy’/exp

OR ‘sentinel lymph node biopsy’ OR ((‘sentinel’/exp OR sentinel)

AND (‘lymph’/exp OR lymph) AND node AND (‘biopsy’/exp OR biopsy))

OR ‘sentinel node biopsy’/exp OR ‘sentinel node biopsy’

OR ((‘sentinel’/exp OR sentinel) AND node AND (‘biopsy’/exp OR biopsy))

OR ‘sentinel node’ OR ((‘sentinel’/exp OR sentinel) AND node))

AND

(‘breast cancer’/exp OR ‘breast cancer’ OR

((‘breast’/exp OR breast) AND (‘cancer’/exp OR cancer))

OR ‘breast neoplasm’/exp OR ‘breast neoplasm’ OR

((‘breast’/exp OR breast) AND (‘neoplasm’/exp OR neoplasm))

OR ‘breast carcinoma’/exp OR ‘breast carcinoma’ OR

((‘breast’/exp OR breast) AND (‘carcinoma’/exp OR carcinoma)))

NOT

(‘lung cancer’/exp OR ‘lung cancer’ OR

((‘lung’/exp OR lung) AND (‘cancer’/exp OR cancer))

OR ‘cervical cancer’/exp OR ‘cervical cancer’ OR

(cervical AND (‘cancer’/exp OR cancer))

OR ‘endometrial cancer’/exp OR ‘endometrial cancer’

OR (endometrial AND (‘cancer’/exp OR cancer)))

AND

[english]/lim AND [humans]/lim

Limiters: Human; English

Publication Date: Last 11 years

### Appendix 2: Data extraction instrument

The general characteristics of all included articles will be recorded and presented in a descriptive table or other format that may include but is not limited to the following^19^:

- Author(s)
- Year of publication
- Source/Origin/country of origin
- Aims/purpose
- Study population and sample size
- Methodology
- Intervention type and comparator
- Concept
- Duration of the intervention
- How outcomes are measured
- Key findings that relate to the RQ

This descriptive table will be separated into two broad categories of interest: ICG and other tracers. It will be piloted with about 15 articles to ensure all relevant results are included and modified if needed.

